# Mutation timing, accumulation and selection in the male germline shape inheritance risk for developmental disorders

**DOI:** 10.64898/2026.04.09.26350474

**Authors:** Matthew D. C. Neville, Sonja Neuser, Rashesh Sanghvi, Joseph Christopher, Kirsty Roberts, Katie Smith, Laura O’Neill, Joseph Hayes, Alex Cagan, Matthew E. Hurles, Anne Goriely, Rami Abou Jamra, Raheleh Rahbari

**Author notes:** Corresponding authors: Matthew D. C. Neville & Raheleh Rahbari.

## Abstract

De novo mutations (DNMs) arising in the parental germline are a major cause of severe developmental disorders. While most DNMs originate in the paternal germline, it remains unclear whether fathers of affected children carry a systematically altered burden of transmissible germline risk, or whether disease largely reflects stochastic outcomes of shared population-wide mutational processes. Here, we combined whole-genome sequencing of 168 parent–child trios with ultra-accurate duplex sequencing of paternal sperm to directly relate transmitted DNMs to the broader mutational and selective landscape of the male germline. In 127 fathers, sperm mutation burden and mutational spectra were indistinguishable from population reference cohorts. Positive selection metrics were likewise concordant, with a global dN/dS of 1.56 (95% CI 1.45–1.67) compared to 1.44 (95% CI 1.17–1.77) in controls and 28 of 32 significantly selected genes overlapping with prior findings. Six fathers harboured a pathogenic early mosaic variant detectable in sperm at allele fractions that ranged from 0.7% to 14.8%. Although these variants generated substantial individual-level risk outliers, they accounted for only ∼11% of the aggregated exome pathogenic burden across the cohort. The remaining burden was distributed across low-VAF mutations, including positively selected driver variants and other rare mutations accumulating with paternal age. Together, these results show that transmissible de novo disease risk is governed primarily by universal germline mutational and selective processes, while early developmental mosaicism produces uncommon but clinically meaningful deviations. This integrated view clarifies how mutation timing, age-associated accumulation and germline selection jointly shape inheritance risk.

## Introduction

De novo mutations (DNMs) are genetic changes that arise during the development of parental gametes and are transmitted to offspring at conception. Most DNMs are single-nucleotide variants (SNVs) or small insertions and deletions (indels). Large whole-genome trio studies show that children inherit ∼50–90 such DNMs, with most of the variation in counts explained by parental age^1–5^. The influence of paternal age is stronger, reflecting a higher mutation accrual rate in spermatogonial stem cells than in oocytes, at roughly 1.5 versus 0.4 mutations per year^4,6^. While the majority of DNMs are benign, pathogenic DNMs account for a substantial fraction of severe paediatric and developmental disorders, with an estimated prevalence of at least 1 in 295 births^7–9^.

From a developmental perspective, DNMs arise from events occurring at different stages of the parental germline lineage, with important consequences for their representation among gametes. Most DNMs arise during the long residence of germline lineages in spermatogonial stem cells or oocytes. Because these mutations occur within extremely large cell populations, any individual event is typically confined to a vanishingly small fraction of gametes, corresponding to very low variant allele fractions (VAFs) in bulk sperm or oocytes. In contrast, a smaller subset of DNMs arise much earlier, during the first weeks of the parents’ own embryonic development. These early events can seed substantial fractions of both somatic and germline tissues, producing mosaic mutations that are detectable in bulk samples at appreciable VAFs, often in the range of ∼1-25%. This distinction between early, higher-VAF mosaic mutations and later, low-VAF age-associated mutations explains their differing transmission properties. In clinical genetics, it has begun to inform emerging approaches to individualized recurrence-risk assessment: when a pathogenic DNM is paternally transmitted and detectable at measurable VAF in sperm, the allele fraction approximates per-conception recurrence risk, whereas absence of detectable mosaicism supports a negligible recurrence risk^10–14^.

Positive selection in the male germline provides a further, mechanistically distinct influence on disease-relevant DNMs. Certain mutations arising in spermatogonial stem cells confer a selective advantage, leading to clonal expansions within the testis and an increasing fraction of sperm carrying these alleles with age. This process, often termed selfish spermatogonial selection, was first inferred from extreme recurrence of specific DNMs in developmental disorder genes and from targeted sequencing of male germ cells^15–17^. Recent advances in duplex sequencing have enabled the systematic quantification of germline selection directly in sperm. Modelling excess nonsynonymous mutations per gene from targeted NanoSeq sperm sequencing identified forty genes under positive selection in spermatogonial stem cells and quantified the age-dependent increase in the fraction of sperm carrying driver mutations^18^. Independent analyses of birth prevalence and DNM datasets show enrichment at many of the same loci, consistent with a shared underlying process linking germline selection, paternal age, and disease risk^19–21^.

Together, these observations point to three broad classes of disease-relevant germline mutations: early embryonic mosaic variants present at elevated VAFs, age-associated mutations present at extremely low VAFs, and positively selected driver mutations that can reach higher frequencies in sperm through clonal expansion. While each of these processes has been studied in isolation, we still lack an integrated view of how they combine to shape the spectrum of transmissible risk in individual fathers. In particular, it remains unclear whether fathers of children with developmental disorders caused by a DNM carry a systematically altered burden or composition of these mutation classes compared to population reference cohorts, or whether most risk reflects shared, population-wide processes.

Here, we address these questions by analysing a developmental disorder trio cohort in which each child’s diagnostic DNM and parental mosaicism status have already been characterised^14^. We combine trio-based whole-genome sequencing with deep, targeted NanoSeq profiling of paternal sperm, enabling direct comparison between mutations detectable in sperm and those transmitted to affected offspring. By integrating mutation timing, variant allele fraction, and selection, and by benchmarking against healthy reference cohorts, we quantify the relative contributions of mosaic, age-associated, and positively selected mutations to disease risk, and assess whether fathers of affected children differ systematically from the general population.

## Results

### Cohort overview and trio DNM counts

We obtained 168 parent-child trio blood samples, each with an affected child carrying a previously identified pathogenic DNM linked to developmental disorder (DD)^14^. Four children carried two pathogenic variants, resulting in 172 diagnostic variants overall. The mean paternal and maternal ages at the time of conception were 35.8 years (range: 23-61) and 32.1 years (range: 22-46), respectively. Whole genome sequencing (median 35x coverage; **Fig 1a**) was performed on all trios and 1 family was excluded following quality control (**Methods**; **Supplementary Table 1**).

**Figure 1.**
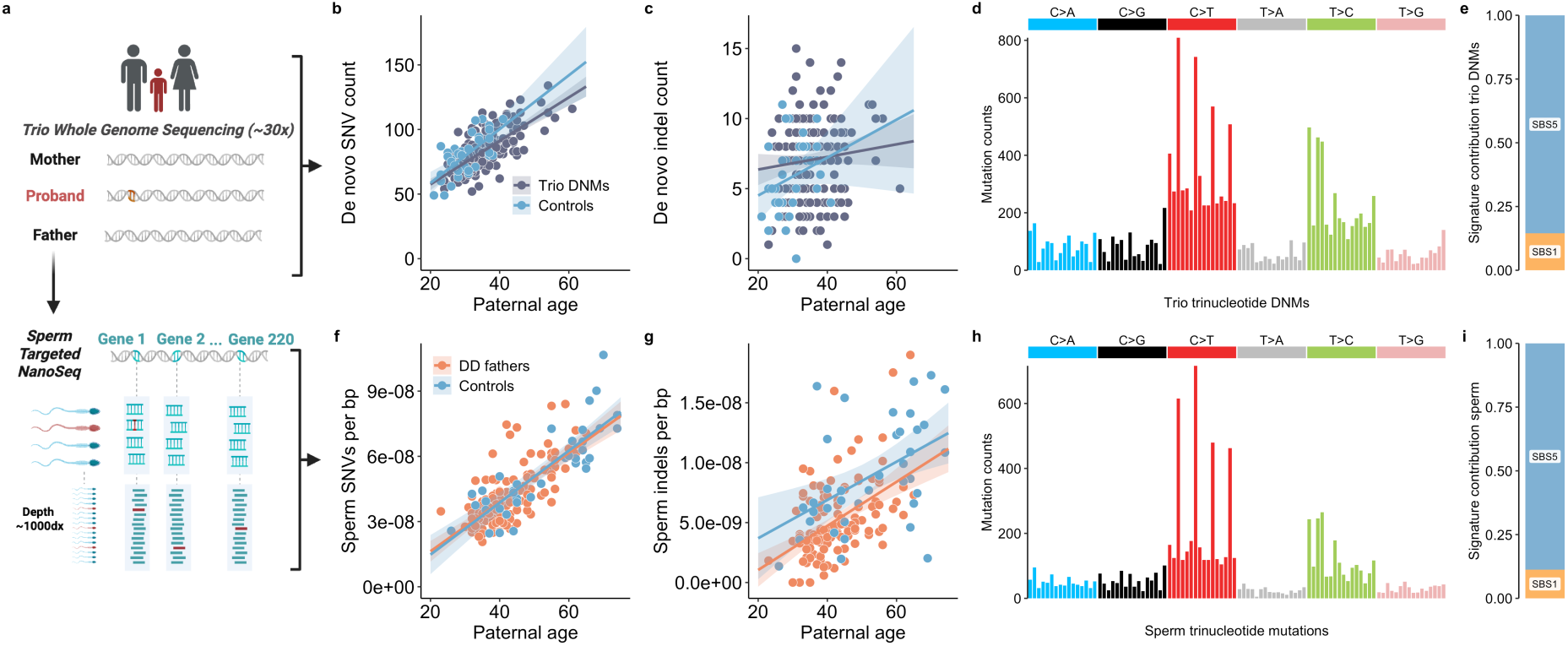
Trio DNMs and sperm mutation burdens and signatures. **a,** Study overview: parent–child trios with DD probands underwent WGS; paternal sperm from fathers was profiled with targeted NanoSeq. Schematic created with BioRender. **b,c,** Transmitted DNM counts vs paternal age for SNVs (**b**) and indel (**c**) mutations in the DD cohort compared to healthy proband controls processed identically. **d**, 96-channel trinucleotide spectrum of trio DNMs. **e**, Contribution of signatures SBS1 and SBS5 to transmitted DNMs in DD trios. **f**,**g**, Sperm SNV (**f**) and indel (**g**) mutations per base pair vs donor age for DD fathers and healthy sperm controls, across the 220-gene capture panel. **h**, 96-channel trinucleotide spectrum of sperm SNVs. **i**, Contribution of signatures SBS1 and SBS5 to sperm SNVs. **b,c,f,g,** Points are mutational load per individual donor; lines are linear mixed-effects regressions with 95% CIs calculated by parametric bootstrapping.

We identified 14,063 single nucleotide variants (SNVs) and 1,181 insertion–deletion (indel) DNMs, restricting analysis to proband variants with allele fraction >0.3 (**Supplementary Table 2**). Of these, 30.5% of DNMs could be phased to the parent of origin, revealing the expected paternal bias (α = 3.16). We find an age-related increase of 1.68 SNVs (95% CI: 1.43-1.94) and 0.045 indels (95% CI: −0.02–0.11) per year of paternal age (**Fig. 1b,c**; **Supplementary Table 3**). Comparing these to a set of 36 control trios not ascertained for disease status of the proband and processed with the same pipeline, we find no significant difference in SNV (p = 0.28) or indel (p = 0.35) accumulation rates. The SNV trinucleotide mutational signatures were inferred to be SBS1 (mean 15%) and SBS5 (mean 85%), the two clock-like signatures consistently observed in DNM and germ cell mutation datasets^6,22^ (**Fig. 1d,e**).

We were able to confirm all previously reported diagnostic DNMs, although 19 required relaxing default filters (e.g. allowing for parental mosaicism or VAF < 0.3 in likely postzygotic mutation cases) or manual inspection (e.g. large or complex indels) (**Methods; Supplementary Table 4**). Beyond the diagnostic variants, additional protein coding DNMs were annotated for potential clinical relevance (**Methods; Supplementary Table 5**). Five variants were classified as pathogenic or likely pathogenic. Two were heterozygous variants in genes associated with autosomal dominant disorders and were returned for clinical review, while three were variants consistent with carrier-only status. The burden of additional reportable variants in DD-associated genes is consistent with a prior large clinical paediatric DD cohort^23^.

### Paternal sperm targeted duplex sequencing

While trio sequencing allows us to identify DNMs transmitted to an affected child, it captures only a single fertilised event. As a result, it provides limited information on how many other parental gametes might have carried the same, or other disease-causing DNMs. We therefore extended our analysis directly to sperm using targeted NanoSeq, which enables accurate detection of mutations from many cells in coding regions for a single sample^24^. Although these approaches differ in multiple ways, three distinctions are especially relevant for interpreting our results. First, trio DNMs capture both maternal and paternal contributions, whereas sperm sequencing only provides access to the paternal component. Second, trio DNMs are subject to ascertainment biases: they derive from fertilised gametes that have successfully developed into embryos and, in this patient cohort, always included individuals with at least one pathogenic mutation identified in the affected child. In contrast, sperm sequencing represents an unbiased sampling of paternal gametes. Third, trio DNMs reflect a single fertilisation event assayed genome-wide, while the targeted sperm sequencing conducted here samples hundreds to thousands of gametes for each father, but only across a restricted set of genomic loci (**Fig. 1a**).

Here, we deep-sequenced 141 semen samples with targeted NanoSeq^24^ from the fathers of the affected offspring (ages 23-67 years), collected a median of 6 years after the child’s birth (range 0-28 years; **Supplementary Table 6**). We applied a 220-gene capture panel designed to include genes (a) recurrently mutated in a large DNM disease cohort^25^, (b) previously linked to positive selection in testis^18,26^, or (c) carrying one or more of the diagnostic DNMs from this cohort (**Supplementary Table 7**). Four samples did not achieve sufficient coverage and ten others failed sample QC filters (**Methods, Supplementary Table 1**), leaving 127 high-quality samples for analysis, 96 of which overlap with the trio DNM sequenced families.

The 127 sperm samples were sequenced to a mean raw depth of ∼6,700× across the panel and, following duplex collapsing, achieved a median duplex coverage (dx) of 951 (IQR 786-1058 dx). This corresponds to sampling ∼950 independent sperm genomes per sample. Variant calling was performed, and multiple quality-control filters were applied, including exclusion of inherited germline variants using a 30% maximum VAF threshold (**Methods**). This resulted in a total of 8,351 SNVs and 1,109 indels called in sperm across the cohort (**Extended Fig. 1a; Supplementary Table 8**). The majority of variants were at extremely low VAFs, with 98.2% being observed in only one cell and only 0.6% reaching VAF >1%, consistent with high polyclonality. We compared the age-related increase in mutations per sperm sample to that of a control cohort of 36 sperm donors previously sequenced with exome NanoSeq^18^. In this, and subsequent analyses, the exome controls were subset to the genome region covered by the 220-gene panel for direct comparisons. Linear regression modelling revealed no significant differences in the age-associated accumulation of mutations between datasets for either SNVs (p *=* 0.66) or indels (p *=* 0.86). The joint slope estimate for SNVs was 1.15 × 10^-9^ mutations per base pair per year (95% CI: 1.00 × 10^-10^ to 1.29 × 10^-9^), and for indels was 1.72 × 10^-10^ mutations per base pair per year (95% CI: 1.24 × 10^-10^ to 2.19 × 10^-10^; **Fig. 1f,g**). The SNV mutational signatures were inferred to be the expected SBS1 (11%) and SBS5 (89%) (**Fig. 1h,i**).

The consistency in both DNM and sperm mutation burdens compared to controls suggests that fathers with a DD child typically have no discernible difference in overall mutation burden in their sperm. However, any paternal-specific enrichment could be obscured by inclusion of fathers whose child’s diagnostic DNM arose maternally or post-zygotically, thereby diluting a signal confined to true paternal transmissions. From the 127 fathers, the origin of the diagnostic DNM in their child was phased as paternal in 43 cases, maternal in 8 cases, and unknown in the remaining 82 (**Methods**). Comparing sperm mutation burdens between individuals with confirmed paternal versus maternal DNM origin revealed no significant differences in either SNV or indel burden (p *=* 0.44 and p *=* 0.64, respectively; **Extended Fig. 2a,b**).

Next, we compared mutation burdens between trio DNMs and sperm for the 96 fathers with both data types available. Sperm substitution burden showed at most a weak association with offspring substitution DNM counts after adjustment for parental age and sequencing-related metrics (p *=* 0.091), explaining little additional variance (56.3% vs. 54.5%). No association was observed for indels (p *=* 0.26). These results align with prior work indicating that parental age explains most variance in DNM burden, and that substantial individual-level deviations are confined to very rare hypermutator ^1,5,27^

### Paternal sperm selection landscape

We next investigated positive selection in the 127 sperm samples by estimating ratios of non-synonymous (N) to synonymous (S) mutations, dN/dS, where a value of 1.0 reflects neutrality. We applied the dNdScv algorithm, which models expected mutation rates while adjusting for trinucleotide context and gene-level covariates known to influence mutability^28^. To improve accuracy in our sperm dataset, we used a previously developed modified version of dNdScv that incorporates base-pair level duplex coverage and CpG methylation levels in testis^18^ (**Extended Fig. 3**).

Across the full cohort, the average dN/dS ratio within the 220-gene panel was 1.56 (95% CI 1.45-1.67), similar to age-matched control samples in the same genomic region (1.44, 95% CI 1.17-1.77) (**Fig. 2a**). To estimate the overall proportion of sperm carrying likely driver mutations in our cohort, we summed the VAFs of known driver mutations in significant positively selected genes per individual. Fitting a quasibinomial regression, we observe a strong positive correlation between age and driver rate (p *=* 4.7e-17) with an estimated 0.35% (95% CI: 0.27%-0.44%) of sperm from individuals at age 30 and 2.0% (95% CI: 1.7%-2.4%) of sperm from individuals aged 60 carrying a known driver mutation, closely matching the age increase in control samples (**Fig. 2b**).

**Figure 2.**
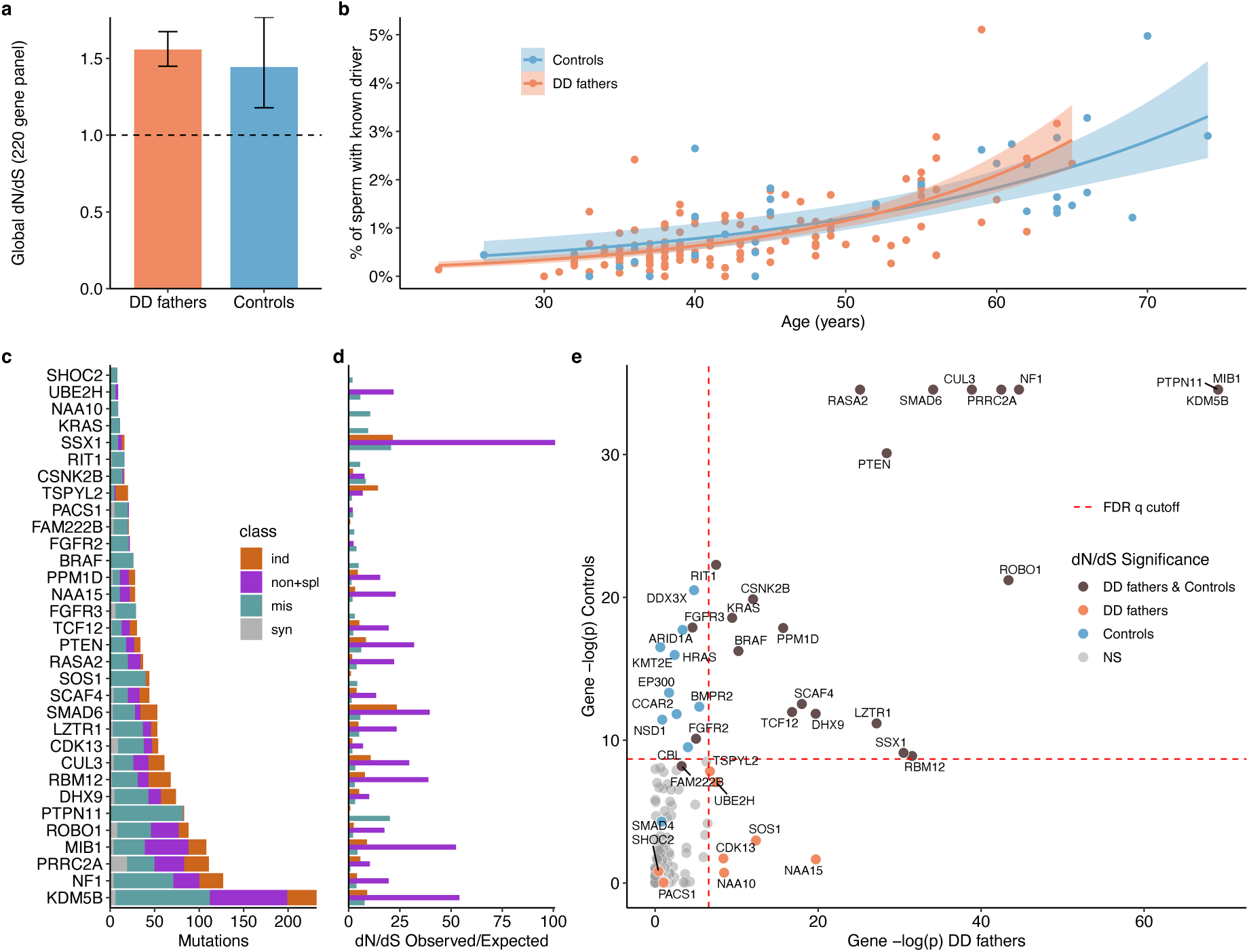
Sperm mutation landscape in fathers of affected children. **a,** Global enrichment of non-synonymous over synonymous mutations (dN/dS) estimated with coverage- and methylation-adjusted dNdScv for DD fathers and controls. **b**, Estimated percentage of sperm per individual carrying a driver mutation by age by cohort. Model fits represent quasibinomial regressions with 95% confidence intervals. **c,** Mutation counts for genes under significant positive selection, split by synonymous, missense, nonsense + splice, and coding indel classes; genes are ordered by total mutation count. **d,** Per-gene, per mutation class dN/dS enrichment; bars coloured as in **c**. **e,** Concordance of gene level dN/dS significance between the fathers of this cohort and controls with p values transformed by negative log. Each point is a gene. Dotted lines indicate the FDR significance threshold in each dataset. Genes that show significance despite not passing the FDR q threshold were significant from hotspot-level tests.

A dN/dS test across the 220-gene panel for the 127 fathers identified 27 genes with significant evidence of positive selection (FDR q < 0.01; **Fig. 2c,d; Supplementary Table 9**). Site-level analysis detected an additional 17 mutational hotspots with ≥3 variants under positive selection (FDR q < 0.01), including 5 in genes not significant at the gene level (**Supplementary Table 10**). In total, of the 32 genes which reached significance, 24 overlap with the previous sperm NanoSeq study18 (*KDM5B, MIB1, PTPN11, NF1, ROBO1, PRRC2A, CUL3, SMAD6, SSX1, RBM12, PTEN, LZTR1, RASA2, DHX9, TCF12, PPM1D, SCAF4, CSNK2B, BRAF, KRAS, RIT1, FGFR3, FGFR2,* and *FAM222B*) (**Fig. 2g**).

Of the remaining eight genes, *SOS1* and *SHOC2* fit the classic profile of RAS–MAPK pathway activators, characterised by recurrent activating missense hotspots that correspond to established pathogenic variants. These same hotspots have previously been implicated in germline selection through targeted testis sequencing^17^ or pathogenic DNM enrichment^21^ respectively. *PACS1* and *CDK13* were also previously identified by Seplyarskiy et al. 2025 through DNM hotspot-level enrichment. While *PACS1*, a protein trafficking gene, has a matching hotspot in sperm and DNMs, *CDK13*, a transcriptional regulator, shows broad loss-of-function enrichment in sperm here without clear correspondence to the reported DNM hotspot. Among the four genes linked for the first time to positive selection in the male germline, *NAA10* and *NAA15*, which are subunits of the same acetylation complex, show mutation patterns consistent with their known dominant developmental disorder mechanisms: clustered missense mutations in *NAA10* and widespread loss-of-function variants in *NAA15*. Finally, *UBE2H* and *TSPYL2* show enrichment for loss-of-function variants and encode proteins involved in ubiquitin-mediated protein turnover and chromatin-associated regulation linked to TGF-β signalling^29^ respectively.

These eight genes which did not previously reach significance in sperm NanoSeq^18^, showed collective dN/dS enrichment in that cohort of 1.89 (95% CI 0.72-4.93), supporting the presence of a true biological signal. Likewise, the 10 previously reported exome-wide significant genes that did not reach significance in this cohort and had sufficient coverage, exhibited collective dN/dS enrichment of 1.27 (95% CI 0.94-1.70). This pattern suggests that their lack of individual significance likely reflects limited power in the opposing datasets rather than absence of selection.

Overall, the strong concordance of magnitude and age correlation of positive selection, as well as substantial overlap of driver genes indicate that the fathers of affected children from this cohort do not exhibit a distinct clonal selection landscape compared to controls.

### Disease causing variants identified in sperm

As the proband diagnostic genes were included in the custom 220-gene NanoSeq panel, 118 diagnostic variant sites compatible with short-read detection and sufficient coverage were interrogated in the corresponding paternal sperm samples. These sites were sequenced to a median duplex depth of 907× (range 201-2735×; **Extended Fig. 4a**), corresponding to per-site 95% VAF exclusion limits ranging from 0.10% to 1.48%.

As these fathers had previously undergone UMI-based single-site testing (∼500× post-collapsing) for parental mosaicism^14^, five diagnostic variants were expected a priori to be detectable in sperm. We replicated all five mosaic variants, with VAFs of 14.8% in *MED13L* (129/873 cells), 14.0% in *SCN1A* (120/738), 2.7% in *NFIX* (27/998), 2.3% in *TUBB2A* (5/220), and 2.3% in *EZH2* (21/927), concordant with the prior UMI-based VAF estimates in the same fathers (**Extended Fig. 4b**). In addition, three of the children’s diagnostic variants that had not previously been detected by UMI-based testing were identified in the corresponding paternal sperm at substantially lower allele fractions: 0.7% in *TRIO* (6/868), 0.15% in *GRIN2A* (1/675), and 0.07% in *SCN1A* (1/1450). The latter two variants fall below conventional mosaic thresholds and are more consistent in allele fraction with age-associated mutations that make up the bulk of detected mutations from NanoSeq. Together, these results demonstrate that deep, panel-based NanoSeq reliably recapitulates known paternal mosaicism and extends detection or exclusion into the sub 1% VAF range.

We also investigated the potential sperm mosaic status of non-diagnostic DNMs identified from the probands. Because these DNMs span the full genome and, unlike the diagnostic variants, were not explicitly included in our target panel, only 17 of 8,439 had coverage in the father’s sperm. Of these 17 variants, only one, an intron variant in *TRPM3*, was detected in the father’s sperm in 3 of 481 cells (0.6%).

To quantify disease-relevant burden beyond the transmitted diagnostic alleles, we defined likely-disease causing mutations observed in sperm as those which are: (i) known ClinVar monoallelic pathogenic/likely pathogenic variants, or (ii) highly damaging variants in high-confidence monoallelic developmental disorder genes^30^ (**Methods**). This framework provides a conservative estimate of pathogenic burden, acknowledging that current gene and variant annotation is incomplete.

We next estimated the percentage of sperm per father carrying a likely disease-causing mutation and extrapolated these values to an exome-wide scale (**Fig. 3**). Across the cohort, most DD fathers had an estimated 1–5% of sperm harbouring such a variant, increasing with paternal age and closely matching controls. A small number of outliers were driven by the six high-VAF pathogenic mosaic variants; removing the mosaic allele returned these individuals to the age-dependent cohort trend.

**Figure 3.**
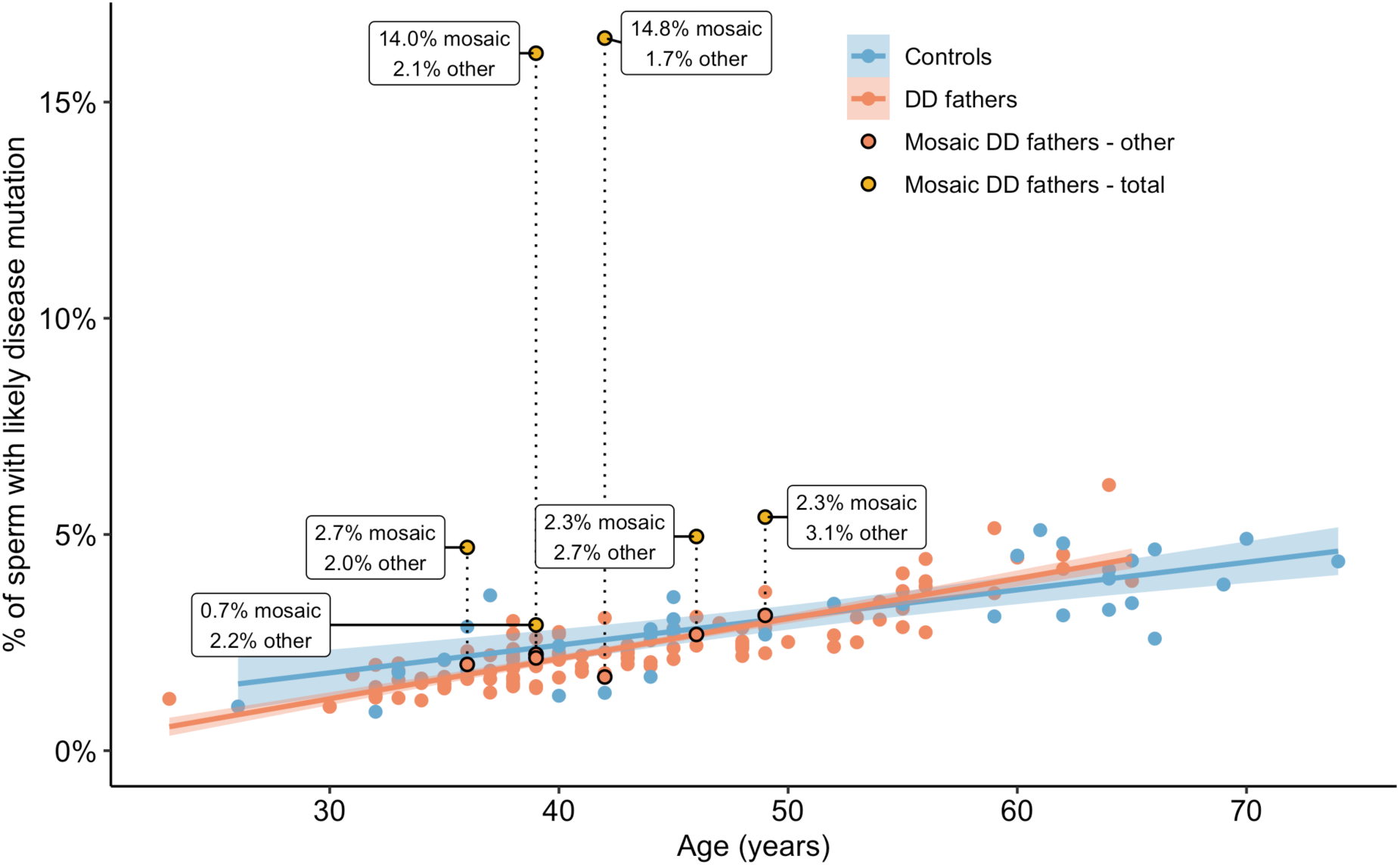
Pathogenic variants in paternal sperm. Estimated percentage of sperm per individual carrying likely disease-causing variants across the exome. For each individual, we summed variant-allele frequencies (VAFs) of variants meeting pathogenic criteria; for DD fathers, estimates were extrapolated from the targeted panel, which directly captured an estimated ∼44% of pathogenic burden, to the whole exome. Groups: controls (n=36; exome-sequenced from Neville et al. 2025), DD fathers without a mosaic pathogenic variant (n=119), and DD fathers with a mosaic pathogenic variant (n=6). For mosaic DD fathers, percentages are shown both excluding (“other”) and including (“total”) the mosaic variant. Model fits are linear regressions with 95% CI bands.

Exome-scale estimates were derived from the 220-gene panel by calibrating to the panel’s disease-mutation capture fraction, which accounts for ∼44.1% of exome-wide pathogenic burden in controls (**Methods**). Importantly, the age-dependent pattern observed within the panel (**Extended Fig. 5a**) was preserved after scaling, with only the absolute magnitude adjusted.

Together, these results suggest two archetypes: fathers whose transmissible risk is driven by age-related mutation accumulation and selection, and fathers with the same background processes plus a fixed contribution from an early embryonic pathogenic mosaic variant.

### Fathers transmitting driver mutations show typical sperm mutation landscapes

In this cohort, it is notable that none of the 13 fathers whose child carried a diagnostic variant annotated as a likely male germline driver had a detectable fraction of the causal allele in bulk sperm at the sequencing depths used here. To replicate this observation, we extended the analysis to an independent cohort of fathers recruited because they had a child with Apert syndrome, a prototypical paternal-age-effect disorder caused by one of two recurrent gain-of-function *FGFR2* mutations under strong positive selection in the male germline^15^.

We sequenced sperm from nine of these Apert fathers using targeted NanoSeq and a gene panel closely matched to that used in the main cohort (**Supplementary Table 7**). One sample failed quality control, leaving eight high-quality samples with a median coverage of 925 dx, yielding 552 SNVs and 70 indels after filtering (**Methods; Extended Data Fig. 1b**). As in the primary cohort, SNV and indel burdens, mutational spectra, selection metrics, and pathogenic-variant burden were all consistent with controls (**Extended Data Fig. 6a-e**). Thus, even in fathers ascertained for transmission of a classic selfish driver mutation, we observed no evidence of a globally perturbed germline mutation landscape.

Despite robust site-specific coverage at the causal *FGFR2* loci (range 364-595 dx; **Extended Data Fig. 4a**), the offspring’s diagnostic *FGFR2* variant was not detected in the sperm of any Apert father by NanoSeq. This absence is expected given prior estimates of *FGFR2* driver variant allele fractions in bulk sperm, including measurements from a subset of these same samples (10^-4^-10^-5^ in Goriely et al. 2003), which are approximately two orders of magnitude below the detection limits achievable at the sequencing depths used here.

Importantly, although the diagnostic driver alleles in the combined 21 fathers from both cohorts were not detected in the corresponding fathers, several of them were observed at low VAFs in sperm from other fathers within the cohort. These included the *PACS1* hotspot (five sperm samples with p.R203W), both *FGFR2* Apert hotspots (3 samples with p.S252W and 2 with p.P253R), and a *PTEN* nonsense variant (one sample with p.R303*). By contrast, despite there being approximately five-fold more diagnostic variants that are not known drivers, only three such variants were observed in another sperm sample from the cohort. This pattern is consistent with germline driver mutations forming a shared, population-wide pool of recurrent high-risk alleles, rather than private expansions confined exclusively to the fathers who transmit them.

### Pathogenic early mosaic variants relative to neutral expectation

An unresolved question is whether fathers of children with developmental disorders are enriched for pathogenic early mosaic variants in sperm relative to the general population. Among the 127 fathers analysed, six harboured a pathogenic mosaic variant detectable in sperm (VAF 0.7-14.8%). When aggregated across the cohort, these variants correspond to a VAF-weighted mean contribution of 0.29% per conception, equivalent to approximately 1 in 341 conceptions (95% CI: 1 in 153 to 1 in 2,755). The breadth of this confidence interval reflects both the small number of pathogenic mosaic events and the skewed distribution of allele fractions, whereby two high-VAF mosaics contribute disproportionately to the aggregate estimate. Additionally, this estimate is restricted to the 220 genes interrogated here and would increase if comparable pathogenic mosaics were present outside the panel.

No pathogenic mosaic variants meeting our criteria were observed exome-wide in the 36 control sperm samples. However, given the rarity of these events and the modest size of the control cohort, we are not powered to formally test enrichment in a case–control framework.

Comparing instead to a prior population-based estimates of mosaic mutation rates in sperm, and shared sibling DNMs^10,11,13,31,32^, normalised to the pathogenicity annotation framework used here, suggests an expected baseline rate between ∼1 in 1,050 and ∼1 in 2010 births (**Methods**). However, the derivation of this estimate depends on assumptions about mosaic mutation rates from different sequencing approaches and mutation model calibration. Consequently, larger, uniformly sequenced cohorts will be required to establish a robust baseline frequency and determine whether pathogenic early mosaicism is enriched in fathers of affected children.

### Relative contributions of mosaic and age-associated germline mutations

Although early mosaic variants generated substantial individual-level transmission risk outliers, they accounted for only 27.5% of the cohort total VAF-scaled pathogenic burden within the 220-gene panel. This reduced to 11.0% of the pathogenic burden when extrapolated to the exome (**Fig. 4d**). The remaining burden comprised low-VAF mutations, including 17.4% likely driver mutations in genes under germline selection (**Methods**) and 71.5% other rare variants.

**Figure 4.**
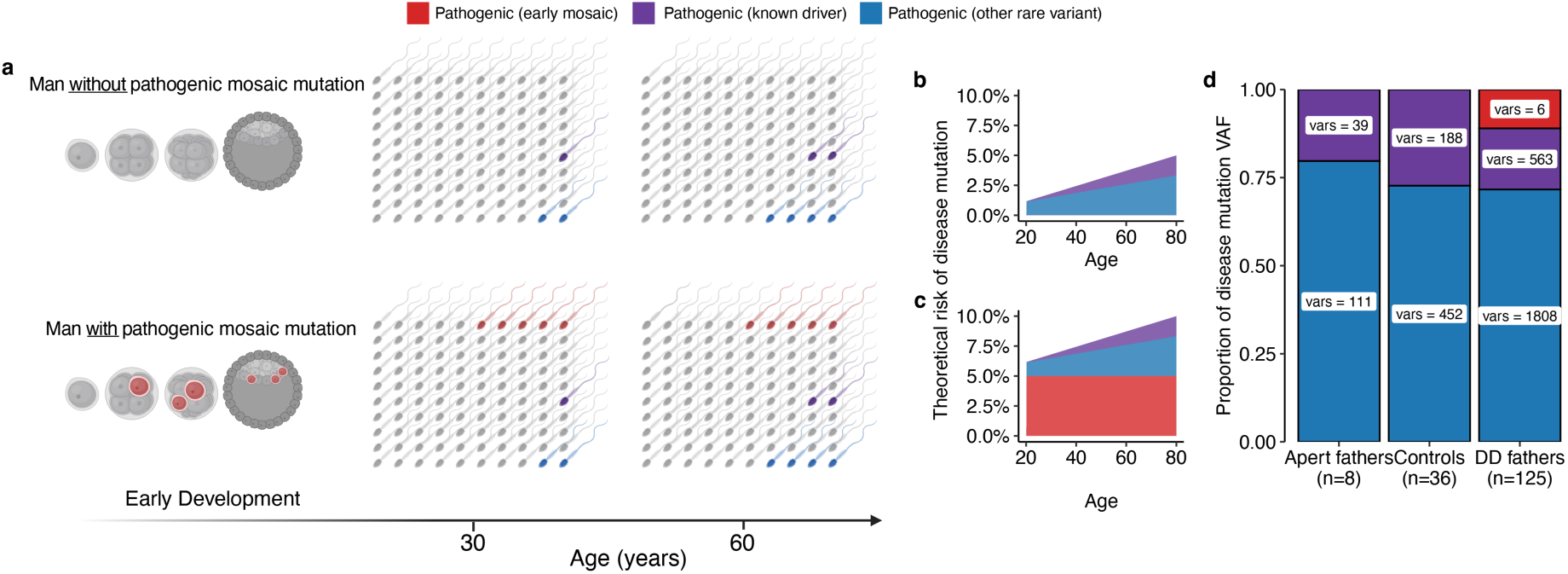
Offspring disease risk from sperm mosaic versus age-related variants. **a-c,** Conceptual model contrasting fathers without (top) and with (bottom) a mosaic pathogenic mutation. Colours denote variant class: mosaic pathogenic allele (red), rare known driver (purple), and other rare variants (blue). Mosaicism creates a time-invariant floor of risk, whereas age-related rare mutations accumulate with age. **a,** Left to right illustrates early development and then 100 randomly sampled sperm at ages 30 and 60 years. Schematic created with BioRender. **b,c,** The expected temporal trajectories of distinct mutation classes in sperm of disease alleles over time. **d,** Cohort-wide contribution of each class to disease-mutation risk after extrapolating to exome for DD and Apert fathers (increasing ‘other rare variant class’ by ∼2.2-fold). Stacked bars show the fraction of the aggregated variant-allele fraction (VAF) attributable to each class in DD fathers (n=127), Apert fathers (n=8), and controls (n=36); numbers inside bars report variant counts contributing to VAF estimate.

These results support a model in which transmissible disease risk arises from three temporally and mechanistically distinct processes (**Fig. 4a-c**). Mutations arising early in parental development can establish a stable fraction of germ cells carrying a pathogenic allele, generating a time-invariant floor of risk within an individual father. In contrast, ongoing age-dependent mutagenesis introduces large numbers of rare variants at extremely low allele fractions. Positive selection acting on a restricted set of driver mutations modestly increases their representation in sperm over time. To illustrate this framework, we modelled the exome-scaled burden of likely disease-causing variants in population reference sperm across paternal age, partitioning the predicted burden into known driver and non-driver components and showing the impact of an added fixed mosaic term (**Fig. 4b,c**). While mosaic variants can dominate risk within a small subset of families, age-associated mutation accumulation and germline selection together account for the majority of pathogenic burden at the cohort level.

## Discussion

De novo mutations are often framed as a stochastic consequence of mutation accumulation during parental ageing. However, disease risk per conception depends not only on how many mutations arise, but also on when they arise and whether particular alleles are amplified by positive selection in the male germline. By integrating trio whole-genome sequencing of 168 families with ultra-accurate duplex sequencing of paternal sperm from 127 fathers of children with developmental disorders, we link pathogenic DNMs transmitted in a single conception to a quantitative readout of their representation across many hundreds of gametes.

At the cohort level, we did not detect evidence that fathers ascertained through an affected child differed from population reference sperm datasets in their overall germline mutation landscape. We observed no cases of germline hypermutation, consistent with prior work showing that extreme elevations in germline mutation burden are rare (<0.1% of trios)^5^. Nor did we observe systematic shifts in the strength or targets of positive selection across the cohort. The global dN/dS across the 220-gene panel (1.56, 95% CI: 1.45-1.67) was comparable to age-matched controls (1.44, 95% CI: 1.17-1.77), and 28 of 32 significant genes overlapped with previous findings^17,18,21^. The four additional new genes reaching significance here (*NAA10, NAA15, UBE2H* and *TSPYL2*) extend the spectrum of known genes likely to be under selection but do not alter its overall landscape. The principal deviations from the age-dependent population baseline were confined to 6 specific early developmental mosaic variants that resulted in single high allele-fraction pathogenic mutations in sperm (range 0.7%-14.8%). Although these mosaics substantially increase transmissible risk within individual families, our study was not powered to determine whether their frequency in the cohort differs from the population baseline, and larger unbiased cohorts will be required to evaluate this directly.

These findings can be understood in terms of three processes that jointly shape paternal pathogenic DNM risk (**Fig. 4a**). First, the timing of mutation acquisition influences transmissibility: mutations arising early in parental development can enter the germline and establish a stable, often measurable fraction of gametes carrying a specific allele. Second, age-dependent mutation accumulation during ongoing spermatogenesis generates a continual influx of new mutations that increase with paternal age but remain at extremely low allele fractions. Third, positive selection acting on a restricted set of driver mutations in spermatogonial stem cells amplifies those alleles through clonal or survival advantages, increasing their representation in sperm with age. Together, this framework explains why early mosaic alleles can dominate risk within a small subset of families, whereas age-associated accumulation and positively selected variants dominate risk at the population level.

Greater resolution sperm sequencing will refine the allele-frequency distribution of low-VAF variants and clarify how different mutational processes contribute to this landscape. At the sequencing depths achieved here, most variants already fall well below levels relevant for transmission of any specific allele, and our data therefore primarily resolve processes influencing aggregate rather than allele-specific risk. Although this depth was insufficient to distinguish low frequency variants arising from clonal selection versus age related mutation accumulation, ultra-deep sequencing has shown that selectively expanded mutations exhibit distinct, higher allele-frequency distributions in sperm^15^. Extending duplex sequencing to ultra-deep depth and incorporating repeat sampling over time should therefore enable separation of these processes and reveal structured allele-frequency distributions shaped by mutation, selection and germline lineage dynamics, without altering the central hierarchy of transmissible risk identified here.

Deep, high-accuracy sequencing of paternal sperm provides a direct quantitative view of the germline mutation landscape at the time of sampling, capturing early mosaic variants as well as the broader pool of age-associated and selectively expanded mutations. However, this view is inherently incomplete. It interrogates only paternal single-nucleotide variants and small indels detectable by short-read methods and does not assess other classes of variation detectable in DNMs^33^ or the maternal contribution. Early embryonic mosaic mutations arise at similar rates in mothers and fathers, reflecting shared early developmental trajectories^2,6,12^. By contrast, oocytes accumulate mutations ∼4-fold slower and are not subject to the same clonal selection pressures observed in spermatogonia. Integrating both parental contributions would therefore modestly increase the age-dependent component of risk while approximately doubling the contribution from early parental mosaics. Within these bounds, deep sequencing of sperm provides a quantitative approximation of the dominant paternal processes shaping transmissible disease risk.

Together, these findings support a model in which transmissible de novo disease risk is governed primarily by universal germline mutational and selective processes, while early developmental mosaicism generates rare but clinically meaningful risk outliers within individual families. In this context, sperm sequencing provides a quantitative framework for interpreting paternal germline mutation burden and situating individual diagnoses within the broader landscape of transmissible risk.

## Data Availability

All processed data underlying the analyses in this study, including variant calls, mutation annotations, and summary statistics, are provided within the manuscript and its Supplementary Information. Additional processed datasets generated during the study are available from the corresponding authors upon reasonable request.
Raw sequencing data (including whole-genome and NanoSeq data) will be deposited in the European Genome-Phenome Archive (EGA) and made available upon publication, subject to controlled access due to participant consent and privacy considerations.

## Extended Figures

**Extended Data Fig. 1.**
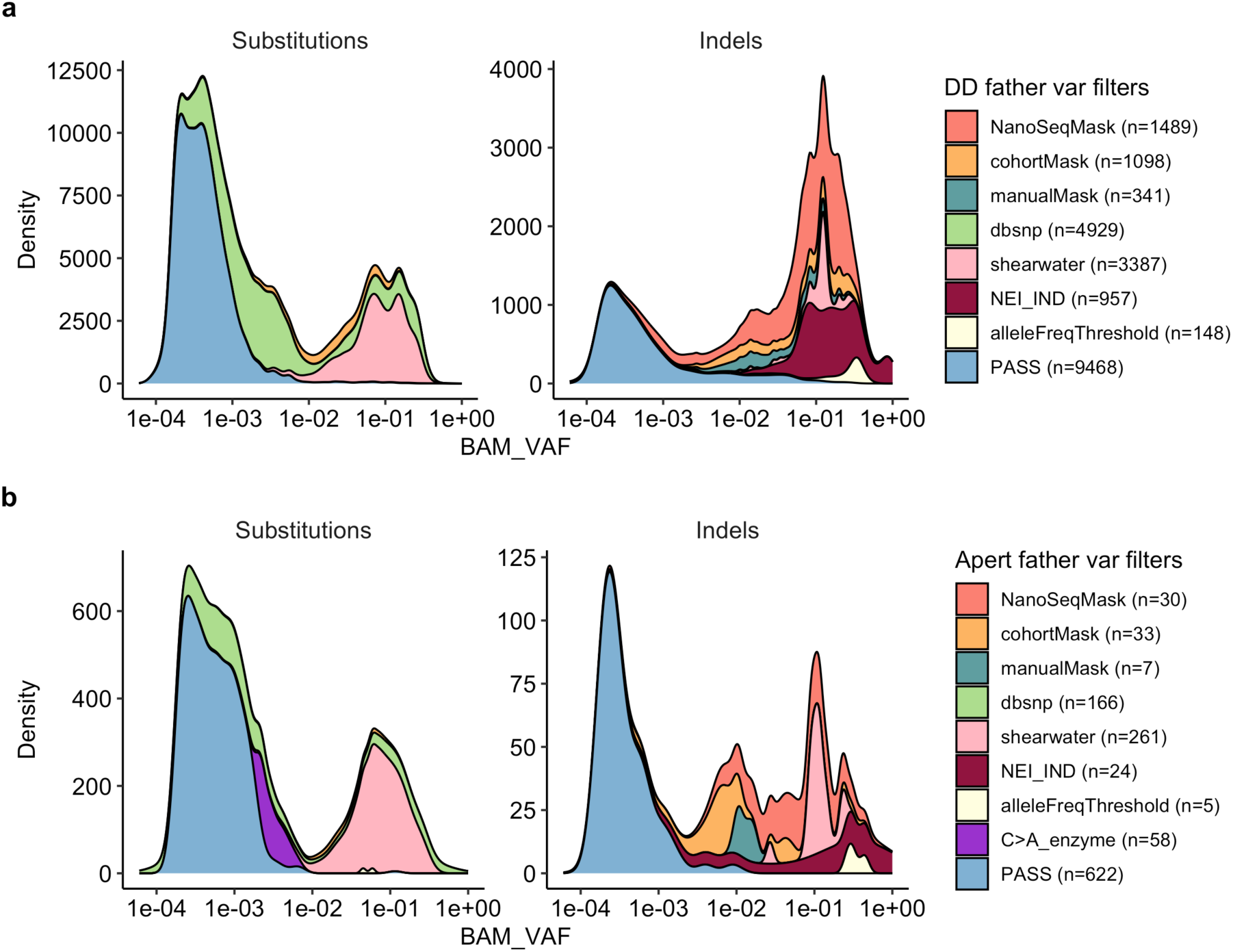
Sperm NanoSeq variant-filtering. **a**,**b**, Variant filtering for fathers of children with developmental disorders (DD) (**a**) and Fathers of children with Apert syndrome (**b**). For each cohort, densities show the distribution of BAM-based variant allele fraction (BAM_VAF; log scale) for substitutions (left) and indels (right). Stacked colours indicate variant subsets defined by the filtering framework, including masked variants from a standard NanoSeq mask, a cohort specific mask, or manual inspection, database polymorphisms (dbSNP), background-error calls (shearwater), indel-specific filters (NEI_IND), an allele-frequency threshold, and an enzyme-specific C> A artefact filter (Methods). “PASS” denotes variants retained after all filters. Numbers in the legends report variant counts per category.

**Extended Data Fig. 2.**
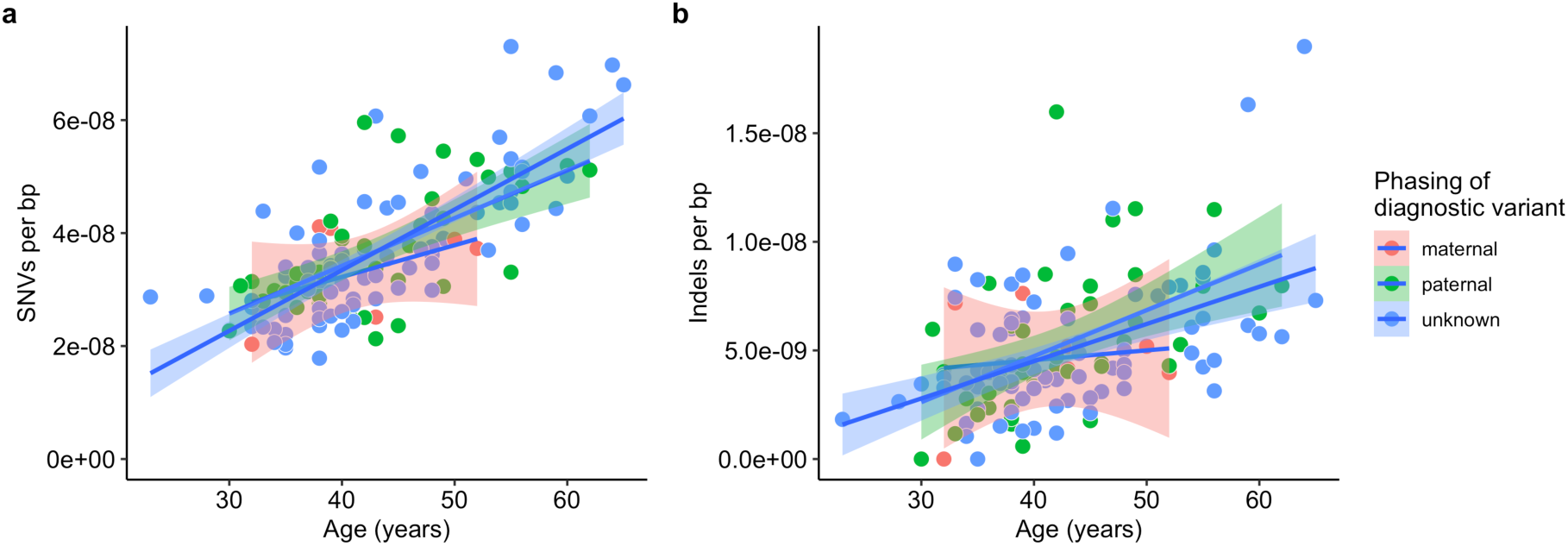
Sperm mutation burdens by phasing of diagnostic variant. **a,b,** Sperm SNV (**a**) and indel (**b**) mutations per base pair versus donor age within the 220-gene target panel, stratified by phasing of the child’s diagnostic variant (paternal, maternal, unknown). Points are individual donors; lines are ordinary least-squares fits with 95% confidence intervals derived from the lm standard errors.

**Extended Data Fig. 3.**
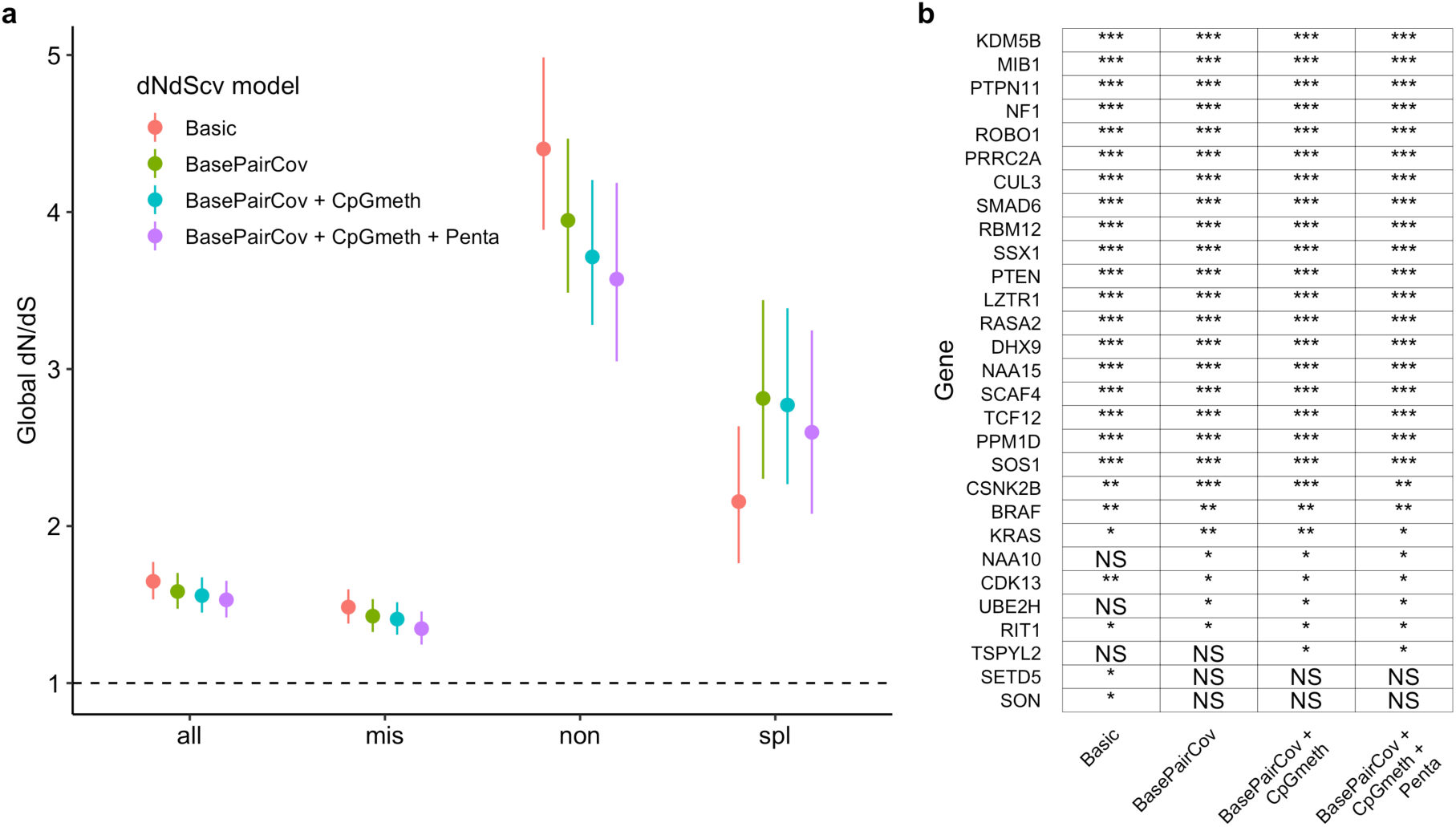
dNdS model selection. **a,** Comparison of global dN/dS values within the gene panel to different modifications to the *dNdScv* algorithm. Categories are all nonsynonymous mutations, missense, nonsense or essential splice. The gene level model includes average gene level duplex coverage but otherwise uses the default parameters such as a trinucleotide mutation model. Additional models show the impact of adding corrections for duplex coverage per base pair (BasePairCov), CpG methylation level (CpGmeth), and pentanucleotide context (Penta). Error bars indicate 95% CIs. **b,** Comparison of per-gene significance in dN/dS tests using the different models. Asterisks indicate significance level of corrected FDR q value : (*q value <0.1, **q value <0.001, ***q value <0.001, NS = not significant).

**Extended Data Fig. 4.**
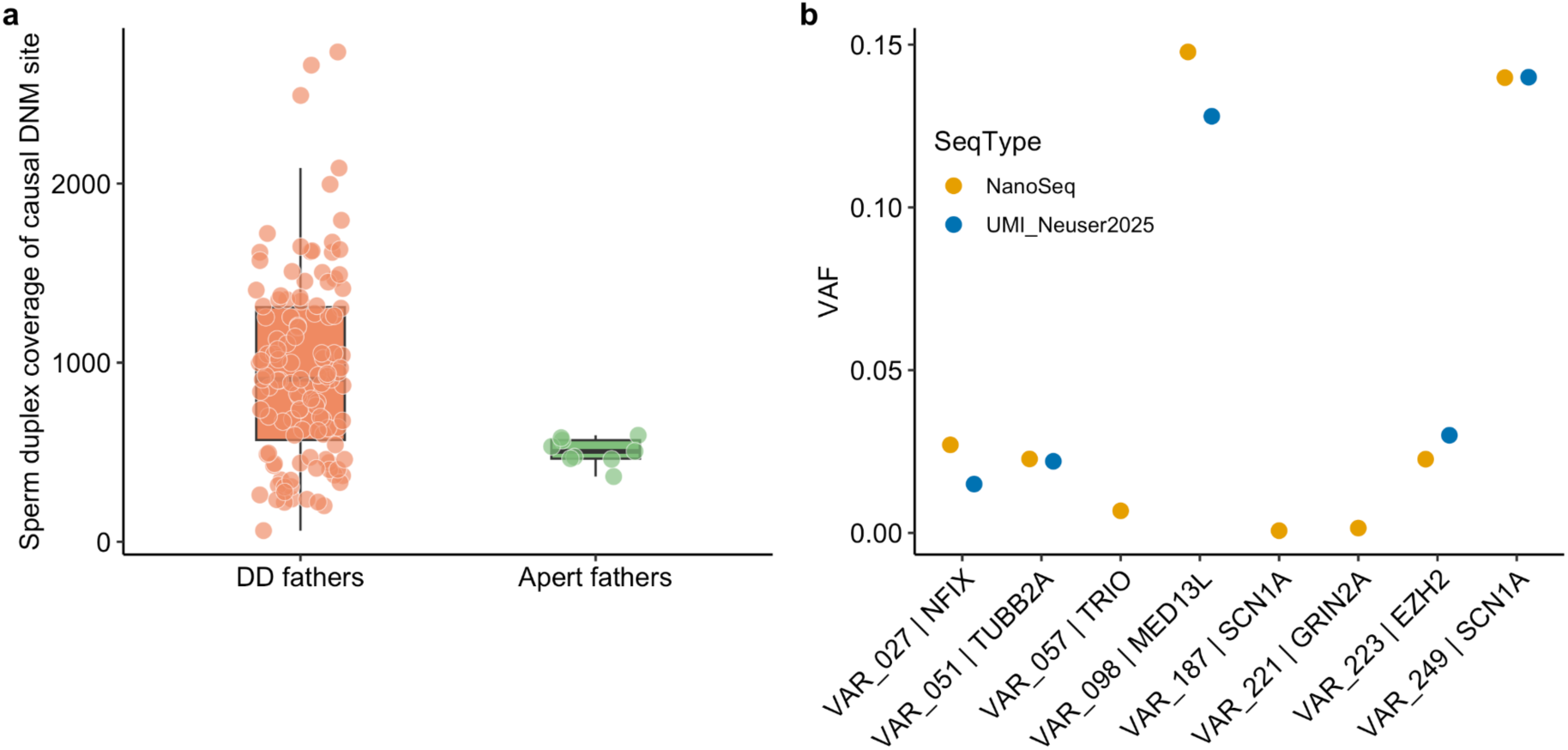
Coverage at causal sites and cross-assay VAF comparison. **a**, Sperm duplex coverage at each proband’s causal DNM site for DD and Apert father cohort. Points are individual samples; boxes show medians and interquartile ranges. **b**, Variant allele fractions for the eight diagnostic variants measured in paternal sperm by NanoSeq and/or by UMI-based sequencing ^14^.

**Extended Data Fig. 5.**
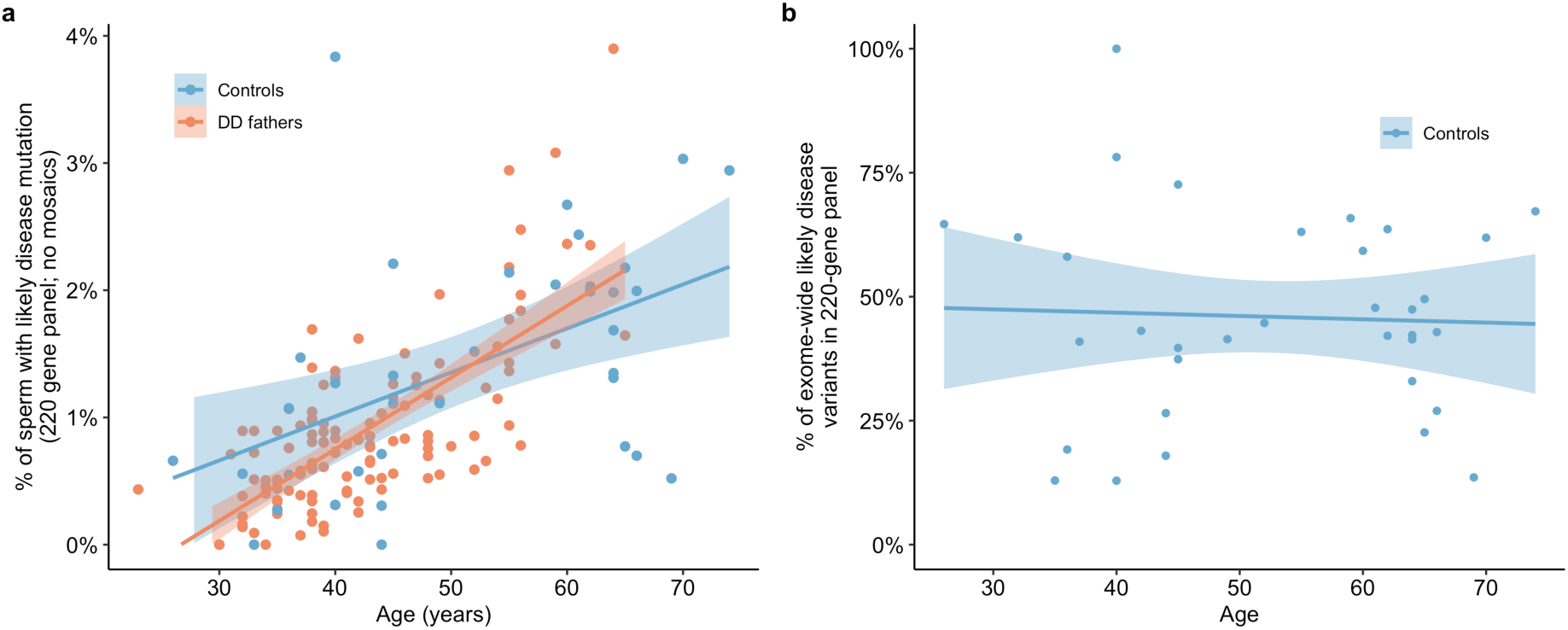
Pathogenic variants in sperm and panel representativeness. **a**, Estimated percentage of sperm carrying likely disease-causing variants within the 220-gene panel versus donor age, for DD fathers (n = 71; excluding 4 mosaic variants) and controls (n = 36). For each individual, the percentage equals the sum of variant-allele fractions of variants meeting pathogenic criteria within the panel. **b**, Percentage of exome-wide likely disease-variant burden for controls that lies within the 220-gene panel versus age. Points are individual donors; lines are linear regressions with 95% confidence intervals.

**Extended Data Fig. 6.**
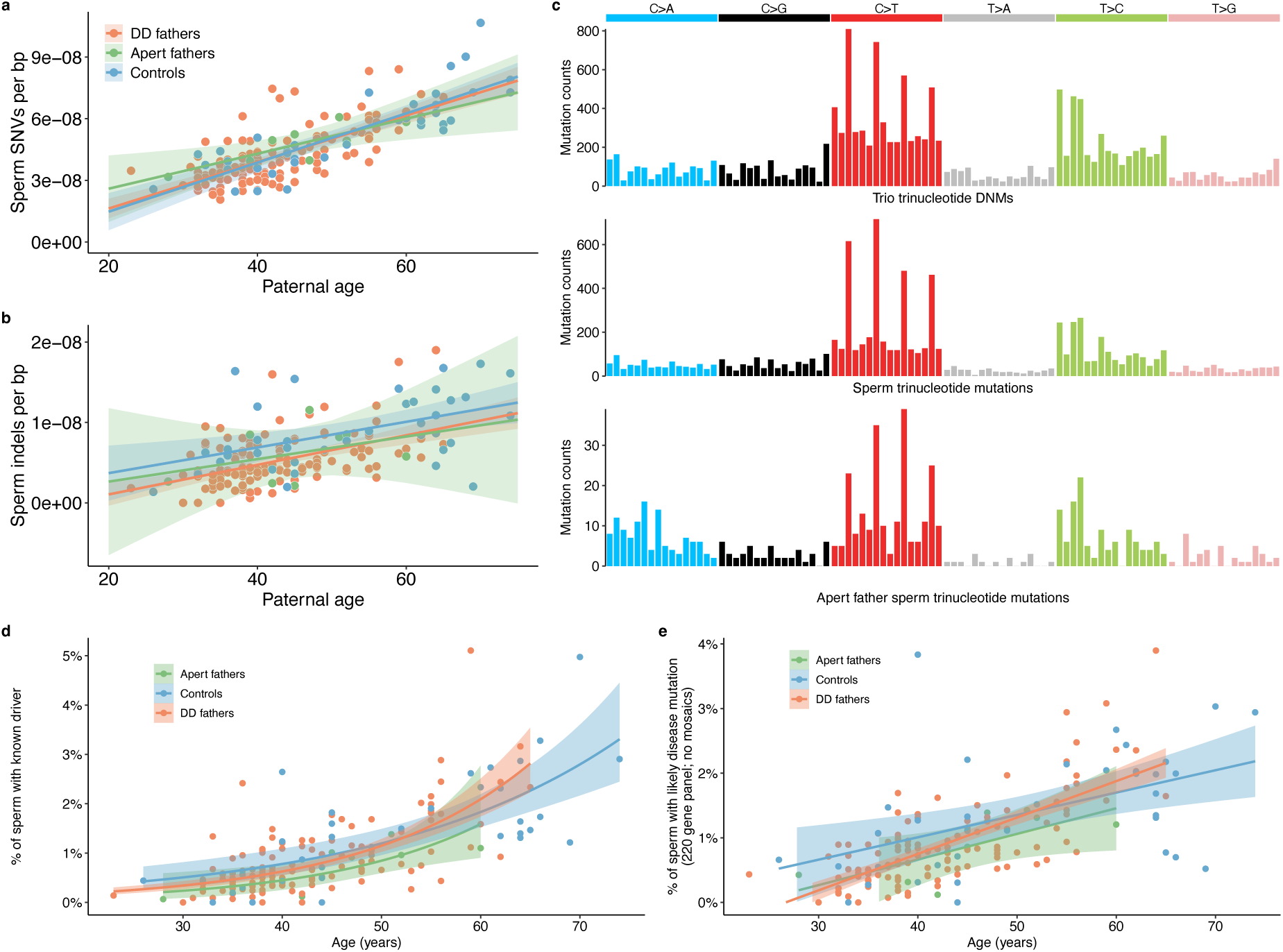
Apert fathers have typical sperm mutation landscape. **a**,**b**, Sperm SNV (**a**) and indel (**b**) mutations per base pair vs donor age for DD fathers (n = 71), Apert fathers (n=8), and controls (n = 36). **c**, 96-channel trinucleotide spectra for the three cohorts. **d**, Estimated percentage of sperm per individual carrying a known driver mutation versus age by cohort. Model fits are quasibinomial regressions with 95% confidence intervals. **e**, Estimated percentage of sperm per individual carrying likely disease-causing variants within the 220-gene panel versus age by cohort; for DD fathers, the four mosaic pathogenic variants are excluded from the sum. **a,b,d,e,** Points are individual donors; lines are linear regressions with 95% confidence intervals.

## Methods

### Ethics

For the primary developmental-disorder trio cohort, the Ethical Committee of the Medical Faculty, Leipzig University, approved genetic testing in a research setting for all probands (014/19-ek). Written informed consent of the parents to publish genetic and clinical data was received and archived by the authors.

Control trios were from 36 trios from 9 healthy families (3-5 children per family) who participated in the Scottish Family Health Study (SFHS). Informed consent was obtained from all participants, and the study was approved by the National Health Service (NHS) East of Scotland Research Ethics Service REC 1 (reference 15/ES/0040).

We obtained single ejaculates from 9 men who had fathered a child with the FGFR2 c.755C>G or c.758C>G variant causing Apert syndrome. All samples were obtained with the permission of the Central Oxford Research Ethics Committee (C03.076).

### Family recruitment and sample collection

Families were recruited from the Institute of Human Genetics in Leipzig, Germany from index cases that underwent next generation sequencing analyses between 2016 and 2021 due to a broad spectrum of disorders. Approximately 300 families were contacted based on inclusion criteria of the index having a diagnostic *de novo* mutation (DNM) and being born in 1996 or later to increase parents’ likelihood of participating.

From the families which participated in the Neuser et al, 2025 study, 168 families had sufficient DNA remaining (>0.2ug) from each member of the trio to perform WGS. Tissue or DNA was obtained from remaining samples previously collected with informed consent. The DNA sources were from peripheral blood collected in EDTA tubes (n = 440), dried blood spots on filter cards (n = 41), saliva (n = 13), buccal swabs (n = 6) or chorionic villus sampling (CVS) (n = 1). There were also 139 semen samples taken from the fathers of the cohort with sufficient DNA remaining for targeted NanoSeq sequencing (>0.3ug).

### DNA extraction

DNA was extracted from EDTA blood, saliva, buccal, and CVS samples using the MagCore 101 Kit. Blood filter cards were cut into small pieces with sterile scissors, incubated in GT buffer, and processed with the MagCore 401 Kit following enzyme digestion. Semen samples were incubated for one hour in GT buffer supplemented with 1 M DTT and Proteinase K prior to DNA extraction using the MagCore 401 Kit. DNA concentration and quality were assessed using a NanoDrop 2000™ or Qubit™ 3.0.

### Whole-genome sequencing and DNM calling

Samples were whole-genome sequenced on the NovaSeq X platform using 150 bp paired-end reads, targeting a mean coverage of 40×. DNM calling was performed using CaVEMan^34^ for single-nucleotide variants, with indels additionally called using Pindel^35^.

Single-nucleotide variants were excluded if they overlapped low-complexity regions, defined as segmental duplications or simple repeats. Sites were removed if more than 10% of reads supported the alternative allele in either parent. Variants with extreme sequencing depth were filtered by excluding sites falling within the top 0.01% of the genome-wide read depth distribution, assuming a Poisson distribution with λ equal to the mean genomic coverage. Variants supported by fewer than two alternative allele reads on each strand were also excluded.

Indels were similarly excluded if located in low-complexity regions. Sites were removed if the parental variant allele fraction was greater than zero, if total depth exceeded 100×, or if coverage in any of the trio members was less than 8×. To reduce strand and alignment artefacts, we retained only variants supported by alternate-allele reads on both forward and reverse strands, and excluded sites where the number of reads classified as unknown or ambiguous was greater than or equal to the number of alternate-allele reads.

Following filtering, candidate de novo mutations were restricted to variants with a variant allele fraction <0.3 to enrich for transmitted germline mutations rather than post-zygotic events. One trio was excluded following quality control owing to an outlier mutation burden that could not be reconciled with the recorded paternal age or with other paternal germline mutation measures, suggesting a metadata inconsistency rather than a biological outlier.

### Diagnostic DNM calling and phasing

Of the 171 diagnostic DNMs in trios that passed quality control, 152 are in the DNM call set generated by the methods and filters described above. Of the remaining 19, fifteen were detected but failed one or more filtering criteria: four had variant allele fractions <0.3, consistent with potential postzygotic origin; seven were present in the unmatched normal panel and/or failed Pindel indel validation filters; two showed evidence of variant reads in a parental sample; one had a low variant-read alignment score; and one had fewer than two supporting reads on one strand. The remaining four diagnostic DNMs were not called, including three variants exceeding the size limits of Pindel and one indel adjacent to an SNV that likely interfered with calling; all four were identifiable by manual inspection in IGV.

Some diagnostic DNMs had previously been phased using long-range PCR followed by nanopore sequencing, as described in Neuser et al., 2025. Additional phasing was performed using read-backed phasing from whole-genome sequencing data using https://github.com/queenjobo/PhaseMyDeNovo, by assigning variants to parental haplotypes based on informative nearby heterozygous sites. Variants were called per sample in gVCF mode using GATK (v4.5.0.0) HaplotypeCaller^36^. gVCFs were merged across each trio, and joint genotyping was performed with GenotypeGVCFs to produce a trio callset used to select heterozygous sites for phasing. Variants were classified as paternal or maternal when phasing was successful using either approach and as unknown when phasing could not be resolved.

### DNM Clinical Annotation

For DNMs called from WGS and not included in the original diagnostic variant list, genes were queried in DECIPHER (v11.34)^37^ to determine any known OMIM^38^ Morbid association and for review of pathogenicity. For variants in genes with an established clinical phenotype, variant-level assertions were retrieved from ClinVar^39^, using entries with ≥1-star evidence. OMIM was reviewed to determine inheritance pattern, clinical synopsis, and relevance to developmental disorders. Heterozygous variants entered on ClinVar as variants of uncertain clinical significance or conflicting interpretation of pathogenicity with an associated autosomal dominant disorder, or an X-linked disorder in a male proband, were re-reviewed using American College of Medical Genetics and Genomics (ACMG) criteria^40^. Variants considered pathogenic or likely pathogenic based on ClinVar entries or manual annotation were then triaged as incidental findings, defined as having potential clinical relevance but unlikely to explain the presenting DD phenotype, or as additional findings plausibly contributory to DD. Variants considered clinically actionable were returned to clinical teams for review.

### NanoSeq libraries and sequencing

Targeted NanoSeq libraries were prepared via sonication and 1 or 2 rounds of pull down of target sequences. They were then PCR amplified and sequenced with NovaSeq (Illumina) platforms to generate 150-bp paired-end reads with 7-8 samples per lane. These steps are described in detail in *Sonication NanoSeq, Library amplification and sequencing, and Hybridization Capture* of Supplementary Note 1 in Lawson et al, 2025^24^.

### NanoSeq base calling and filtering

Sperm samples were processed using the NanoSeq calling pipeline (https://github.com/cancerit/NanoSeq). BWA-MEM^41^ was used to align all sequences to the human reference genome (NCBI build37). We leveraged the high sequencing depth and high polyclonality to exclude variants with BAM VAF > 30% instead of using a matched normal.

Default parameters of the calling and filtering pipeline for targeted NanoSeq were used. These included masking common germline variants from dbSNP^42^ and common NanoSeq artefacts described previously^43^. Post-calling filters were applied to define the final PASS set. Variants were reassigned to alleleFreqThreshold if the duplex VAF ≥ 0.30, or if duplex consensus coverage ≤ 11. A cohort-level mask (cohortMask) removed loci that were excluded by default filters in 2 or more samples in the cohort. A list of 17 recurrent artefacts identified by inspection was marked manualAnnot. For the Apert father samples, an additional filter (C>A_enzyme) excluded a small number of high VAF (BAM_VAF > 0.002) C>A and G>T substitutions, due to an error profile caused by a mistakenly substituted enzyme. The resulting VAF distributions of passed variants concentrate at low VAFs with a small tail of higher VAF variants as expected (**Extended Fig. 1**).

### Sperm sample filtering

We sequenced 141 stored sperm DNA aliquots from fathers in the DD cohort with > 200ng of DNA remaining. After sequencing, 4 were excluded for insufficient depth (<150dx), all from low-input samples (<700 ng). We received 9 semen samples from fathers of children with Apert syndrome; all met input requirements and achieved sufficient coverage for analysis.

To guard against contamination, we examined sequencing data for (i) exogenous DNA and (ii) non-sperm cell DNA. Exogenous contamination was assessed using the ratio of masked (SNP/noise filters) to pass variant calls across the panel; 4 samples with a masked:pass ratio >5 were excluded. Suspected somatic contamination was defined by an outlier mutation burden together with multiple canonical clonal-hematopoiesis driver calls, as blood is the most commonly found contaminating cell in semen. Six primary cohort samples and one Apert father sample were excluded from this filter. This left 127 primary cohort and 8 Apert father semen samples for downstream analyses.

### Mutation burden regressions

For sperm analyses, corrected burdens were derived from the raw metric (duplex-supported variant calls divided by duplex-callable bases) with two adjustments. First, to limit inflation from germline selection genes, variant and coverage counts were restricted to the 220-gene panel after excluding coding bases of 45 germline driver genes identified across this study and the Neville et al., 2025 study. This restriction was applied uniformly to DD fathers, control exomes, and the Apert cohort. Second, burdens were computed using only variants with VAF < 1%, because rare high-VAF events >1% are sparse yet disproportionately elevate per-sample burden on a targeted panel.

Linear regression was used to estimate age-associated accumulation and to compare burdens between datasets. For SNVs and indels separately, we fit a null model with age and dataset as additive fixed effects and a full model with an age×dataset interaction to test slope differences. Models were fit with lm() in R and compared with anova(). Significance was assessed at P < 0.05.

For plotting, 95% confidence intervals around fitted lines were obtained by nonparametric bootstrap. We drew 1,000 bootstrap resamples, refit the model in each, and generated predictions across a 5-year age grid spanning the observed range. The 2.5th–97.5th percentiles of the bootstrap predictions at each age define the confidence bands.

### Comparison of mutation burdens

To evaluate the relationship between paternal sperm mutation burden and offspring de novo mutations (DNMs), we restricted analysis to the 96 fathers for whom both sperm sequencing and trio-based DNM counts were available. We then used multiple linear regression to test whether sperm burden predicted offspring DNM counts after adjusting for paternal age (PatAge), maternal age (MatAge), the median sequencing coverage of each of the 3 trio genomes (QC), the median DNM VAF (QC_4_) and the age difference between sperm sampling and conception (AgeGap). For each mutation class (SNVs and indels), we fit two nested models:

**Base model:**

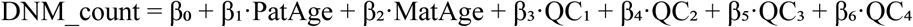

**Full model:**

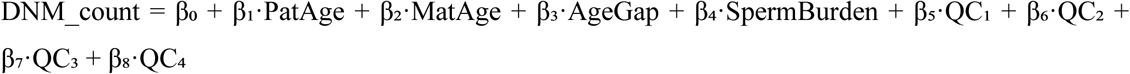

The increase in unadjusted R² between the base and full models was used to quantify the additional variance explained by sperm burden. All regression analyses were performed in R using the lm function.

### Mutational signature analysis

Mutational signatures were extracted based on non-negative matrix factorization using SigProfiler (https://github.com/AlexandrovLab) using SigprofilerExtractor 1.1.24. To account for differences in the trinucleotide background between targeted sequenced sperm samples and known mutational signatures, the mutational matrix was normalised before extracting signatures. This normalisation, similar to the mutation burden adjustment, was accomplished by multiplying the 96 categories of trinucleotide mutations by their ratio relative to the full callable genome trinucleotide contexts. These normalised values were then rounded to the nearest integer and used as the input matrix for SigProfiler. Signatures were deconvoluted into known COSMIC v3.4 signatures^44^.

### Quantifying selection with dN/dS

To examine genes under positive selection and quantify global selection we used the *dNdScv* algorithm^28^. Analyses were restricted to the 220-gene targeted panel. Sites and their genes were considered under positive selection if a site reached an FDR q-value < 0.01. One additional gene, *MECP2*, showed significant enrichment of missense and loss-of-function variants at the gene-wide level but was excluded due to significant enrichment of synonymous mutations (observed/expected = 16.0; q = 1.4e-09) and a non-significant local dN/dS value (q = 0.15).

The *dNdScv* algorithm was extended using base pair level duplex coverage and methylation level. These model extensions are described briefly here and in detail in Supplementary Note 3 of Neville et al 2025. The base pair coverage correction was implemented by multiplying the mutational opportunities matrix by not just the number of base pairs in that context but by the exact number of duplex molecules with sequencing coverage in that context across the cohort. The methylation level adjustment involved extending the mutational opportunities matrix from 192 contexts (96 trinucleotides + transcribed vs non-transcribed strand) to 208 contexts, where each of the 8 possible CpG trinucleotide opportunities was split into 3 contexts based on percent methylation at that site in testis: low (0-10% methylation), mid (11-40% OR unknown), and high (>40% methylation) methylation.

We did not use a pentanucleotide extension because applying tri- versus pentanucleotide models yielded no changes in gene-level significance and no significant differences in context-specific rates (**Extended Fig. 3**). Significant difference in rates was evaluated two-tailed z-test was performed for each tri- to pentanucleotide comparison, adjusted for multiple testing using the Benjamini-Hochberg procedure.

The global dN/dS comparison used all DD father samples but restricted the control sample set to samples within the age range of the DD fathers. This was performed to avoid a bias in overall selection levels due to the dependence of selection level on age.

To evaluate selection at hotspots we applied a modified version of the *dNdScv* sitedNdS function^45^ that detects positive selection at specific base pair sites. We tested each coding position with ≥3 observed mutations against its trinucleotide context- and coverage-adjusted expectation using the dNdScv site test. The coverage-adjusted expectation was generated similarly to the gene level test by using a dN/dS model fit with base pair level correction and adjusting for the duplex coverage of each recurrently observed mutation tested.

### Variant annotation

DNMs and sperm variants were annotated using Ensembl’s Variant Effect Predictor (VEP)^46^ with added custom annotations of mutation context, ClinVar release 2025.07.29^39^, Combined Annotation Dependent Depletion (CADD) version GRCh37-v1.7^47^ and average methylation level in testis. Methylation data was obtained from whole genome shotgun bisulfite sequencing methylation data from two male testes samples (ENCFF638QVP & ENCFF715DMX) from the ENCODE project^48^. The average methylation level was calculated by selecting CpG sites with coverage of 3 or more and averaging the percent of sites methylated between the two samples.

Using these annotations variants were annotated as one of two non-mutatually exclusive labels: “likely disease” or “likely driver”. Likely disease mutations met at least one of two criteria: 1) Reported in ClinVar as pathogenic, likely pathogenic, or reported as ‘Conflicting_classifications_of_pathogenicity’ where the conflict was between reports of pathogenic/likely pathogenic and ‘Uncertain_significance’ with no reports of benign or likely benign and not specified as a recessive condition or 2) Were a highly damaging variant in a high confidence monoallelic developmental disorder genes from DDG2P^30^. More specifically the DDG2P genes met all three of the following criteria: a) allelic requirement that was not “biallelic_autosomal” b) confidence being “strong” or “definitive” and c) a molecular mechanism of “loss of function”. Highly damaging was defined as being a ‘HIGH’ impact variant in VEP annotation (frameshift splice_acceptor, splice_donor, start_lost, stop_gained, or stop_lost) or a missense variant with CADD^47^ score >30 (top 0.1% damaging) and gnomADe_AF < 5e-6.

Variants were defined as a likely driver if they met the ‘highly damaging’ criteria defined above in one of the 36 significant germline selection genes with loss-of-function mutation enrichment (ptrunc_cv < 0.1 or pind_cv < 0.1) here or in Neville et al, 2025, or if they were in one of the 30 significant mutation hotspots from this paper or Neville et al, 2025.

Variants were defined as mosaic in sperm if they were called in > 2 duplex molecules and above 0.5% VAF.

### Cell fraction estimates

To estimate the percentage of sperm per individual carrying a likely driver or likely disease mutation we summed the duplex VAFs of all variants in that class. For example, if an individual had three driver mutations each observed once with a duplex coverage of 100 at each of those sites, each of those variants would have a duplex VAF of 1/100 = 0.01. The sum of VAFs in this example would then be 0.03. At low fractions such as 0.03, the mean count per cell is approximately equivalent to the percentage of sperm with this mutation class (3%) and thus the driver and disease mutations are reported as percentage estimates. Note that we cannot exclude the possibility that some of these mutations are from the same cell.

To generate Fig. 4b,c, we fit linear models of exome-scaled VAF-summed burden versus age in the exome control sperm samples, separately for likely driver variants that are also likely disease and for all likely disease variants. The driver component was taken directly from the age-dependent fit of known drivers. The non-driver component was calculated as the age-specific difference between total likely disease burden and driver burden. Predicted values across an age grid were visualised as stacked area plots. An additional fixed mosaic term was added to illustrate the impact of early developmental mosaicism on overall risk.

### Extrapolating from panel to exome

We estimated per-donor pathogenic burden as the percentage of sperm carrying a likely disease mutation, computed as the sum of VAFs within the 220-gene panel as described. To place these panel measurements on an exome scale, we calibrated a panel capture fraction in controls that had both panel-restricted and exome-wide estimates. For each control, we calculated the ratio of panel burden to exome burden and tested its dependence on age using ordinary least squares; no age effect was detected (p = 0.81; **Extended Fig. 5b**). We therefore used a single capture fraction, of 0.441, representing the proportion of exome-wide pathogenic burden contained in the 220-gene panel. This adjustment is supported by the finding that panel and exome burdens were strongly correlated across individuals (p = 3.2 × 10^-7^). Exome-wide percentage estimates for DD fathers were obtained by scaling the panel burden. Where counts were required, we applied the same factor to the panel count and rounded to the nearest integer.

### Estimation of baseline paternal sperm mosaic risk

For the cohort-level estimate of pathogenic mosaic burden in fathers of affected children, we calculated the mean VAF across all 127 fathers and converted this to a per-conception probability. Ninety-five percent confidence intervals were obtained by nonparametric bootstrap (10,000 resamples).

To estimate a population baseline rate of pathogenic paternal sperm mosaicism, we synthesised four published deep-sequencing studies of sperm and one study of shared DNMs in large families^10,11,13,31,32^. Across these datasets, men carried approximately 2–4 detectable sperm mosaic SNVs per individual at VAF ≥1%. Given heterogeneity in sequencing depth, variant calling thresholds, and mosaic definitions across studies, we used this range (2–4 events per individual) to define lower and upper baseline bounds.

We decomposed baseline pathogenic mosaic risk into three parameters: (a) the number of detectable paternal sperm mosaics per individual (2–4), (b) the fraction of WGS callable genome represented by our exome panel region (∼2.8%), and (c) the probability that an exonic SNV meets our likely-disease criteria, estimated from our coding mutation model as approximately 1 in 4,200 genome-wide detectable SNVs. The expected per-conception probability of a detectable paternal sperm mosaic classified as likely disease-causing was therefore approximated as: risk ≈ (2 or 4) × 0.028 × (1 / 4,200) ≈ 9.5 × 10^-4^, yielding a baseline rate between ∼1 in 2,010 and ∼1 in 1,050 conceptions.

## Declaration of generative AI and AI-assisted technologies in the writing process

During the preparation of this work the authors used ChatGPT in order to improve the readability of the manuscript. After using this tool, the authors reviewed and edited the content as needed and take full responsibility for the content of the published article.

## Data availability

Raw sequencing data will be uploaded to the European Genome–Phenome Archive upon publication.

## Code availability

All scripts are available on github at https://github.com/mattnev17/sperm-nanoseq-dd-fathers.

## Acknowledgements

We thank all participating families and sperm donors. We thank Katrina Andrews and Joanna Kaplanis for helpful suggestions on the manuscript. This research is supported by core funding from Wellcome Trust grant 220540/Z/20/A. R.R. is funded by Cancer Research UK (C66259/A27114) and Medical Research Council (MR/W025353/1). This study was supported by the Deutsche Forschungsgemeinschaft (537144118537144118 with AB393/9-1 to Rami Abou Jamra and NE2706/2-1 to Sonja Neuser). We acknowledge funding support from Wellcome (219476/Z/19/Z) and the National Institute for Health Research (NIHR) Oxford Biomedical Research Centre Programme to AG. JC was supported by the NIHR Cambridge Biomedical Research Centre (NIHR203312).

## Author Contributions

R.R. conceived and designed the study and, together with M.E.H., supervised the project. S.N. and R.A.J. collected and prepared all samples and participant data from the primary cohort. A.G. collected and prepared all samples and participant data from the Apert father cohort. K.R., L.O., J.H., and A.C. contributed to sample extraction and sequencing implementation. M.D.C.N. led analysis of sperm data, R.S. led DNM analyses, and J.C. clinically annotated DNMs. M.D.C.N. and R.R. wrote the manuscript, and all authors reviewed and edited the final version.

